# *Candida (Diutina)* mesorugosa in Non-Albicans Candida Species Clinical Isolates in South West Nigeria

**DOI:** 10.1101/2020.07.21.20157370

**Authors:** Oloche Owoicho, Judith Uche Oguzie, Tosin Segun Ogunbiyi, Toyin Adulsalam, Philomena Eromon, Onikepe Folarin, Ernest Uzodimma Durugbo, Christian T. Happi

## Abstract

**Introduction:** The emerging role of non-albicans *Candida* species (NACS) as causal agents of mild to life-threatening mycoses is increasingly being appreciated. Some NACS are known for intrinsic resistance or reduced susceptibility to some antifungal drugs. To inform on therapeutic options and management of candidiasis, we embarked on *Candida* species surveillance in South west Nigeria.

**Method:** We obtained retrospective yeast isolates from urogenital samples of patients in a tertiary hospital in South west Nigeria. Among 47 isolates identified phenotypically, we encountered a yeast which failed to produce pseudohyphae in human serum but was morphologically similar to *Candida albicans*. We characterized this yeast by sequencing the internal transcribed spacer (ITS1-5.8S-ITS2) region of the ribosomal DNA.

**Results:** A BLAST search and sequence homology identified the yeast as *Candida* (*Diutina*)*mesorugosa*, with a strong bootstrap.

**Conclusion:** *Candida mesorugosa*, a rarely isolated yeast from clinical samples worldwide, should be considered among potentially relevant NACS in Nigerian clinics. To the best our knowledge, this is the first report of *Candida mesorugosa* from a clinical sample in Nigeria. This finding confirms the need for more specific methods, such as DNA sequencing, for extensive surveillance of NACS.

## Introduction

*Candida* (*Diutina*) *mesorugosaCandida* (*Diutina*) *rugosa, Candida* (*Diutina*) *pseudorugosa* and *Candida* (*Diutina*) *neurorugosa are members of Candida rugosa* complex (Paredes et al. 2012). Members of *Candida rugosa* complex exist predominantly in the environment, and are widely used in the food, cosmetic and pharmaceutical industries for their lipase (Benjamin and Pandey 1998). However, *C. rugosa* complex are associated with mastitis in dairy cattle (Scaccabarozzi et al. 2011). Episodes of human candidemia caused by *C. rugosa* complex have also been reported (Pfaller et al. 2010; Singh et al. 2011). Like many non-albicans *Candida* species (NACS), reduced susceptibility to azoles was observed in the *rugosa* complex (Pfaller et al. 2010). Taken together, *C. rugosa* complex are considered emerging agents of antifungal refractory infections.

Surveillance of *Candida* species has been recommended due to increasing threat of NACS and to inform on strategies for prevention and management of recalcitrant mycoses. However, in under-resourced countries, including Nigeria, surveillance is hampered by use of less robust and sensitive techniques. To circumvent this challenge, we collected retrospective yeast isolates recovered from patients, with the aim of speciating the yeasts, using molecular diagnostics, DNA sequencing and Bioinformatics approaches. This short communication is part of the ongoing project.

## Materials and Methods

Forty-seven putative yeast isolates recovered from urogenital samples of patients from February to May 2018 were obtained from Ladoke Akintola University of Technology Teaching Hospital, Osogbo, a tertiary hospital in South west Nigeria. The isolates were sub-cultured on Sabouraud dextrose agar at 37°C for 48 hours and initially identified using CHROMagar(tm) Candida (ChromAgar, Paris, France). An isolate, here designated NG06, which did not produce pseudohyphae in human serum, but was morphologically similar to *C. albicans* was selected for molecular identification. The isolate was subjected to heat shock (da Silva et al. 2012) then genomic DNA was extracted using Qiagen DNeasy Blood and Tissue Kit (Qiagen, Germany) following the manufacturer’s instructions. ITS1-5.8S-ITS2 region of the rDNA was amplified by PCR using ITS1 (5’-TCC GTA GGT GAA CCT GCGG-3’) and ITS4 (5’-TCC TCC GCT TAT TGA TAT GC-3) universal primers (Integrated DNA Technology, USA). The PCR reaction mixture comprised 5 µL of DNA template, 0.4 µL of each primer and 12.5 µL of One Taq Quick Load Master Mix (2X) (New England BioLabs) made up to a final volume of 25 μL by PCR grade water. PCR amplification was performed on Mastercycler Pro-Vapo. project (Eppendorf AG,Hamburg, Germany) using the thermo-cycling conditions described previously (Solimani et al. 2014). The amplicons were subjected to 2% agarose gel electrophoresis and visualized. The amplicons were sequenced on Applied Biosystems 3500xL Genetic Analyzer (Thermo Fisher Scientific, UK). For bioinformatics analyses, the sequence was imported to Unipro-UGENE, viewed and edited where necessary. Further, the sequence was compared against a non-redundant database (NCBI) to infer homology based on the hits found. A multiple alignment was generated by combining the sequence and the best hits. The best hits were chosen based on bits score, e-value, maximum score and query coverage. Phylogenetic trees were generated from the alignment dataset, producing a Maximum-likelihood tree which was estimated by using Kimura’s two-parameter method implemented in Unipro-UGENE version 34.0. The reliability of the phylogenetic tree was assessed by 1000 bootstrap resampling. The sequence was submitted to NCBI (GenBank accession no. **<u>MT548606</u>**).

## Results

The PCR amplification yielded nucleic acid product of 390bp (Fig. 1). The BLAST search identified the sequence as *C*. (*Diutina*) *mesorugosa*, which clustered with known sequences of *Diutina mesorugosa*, with strong bootstrap support (Fig.2).

**Figure 1.**
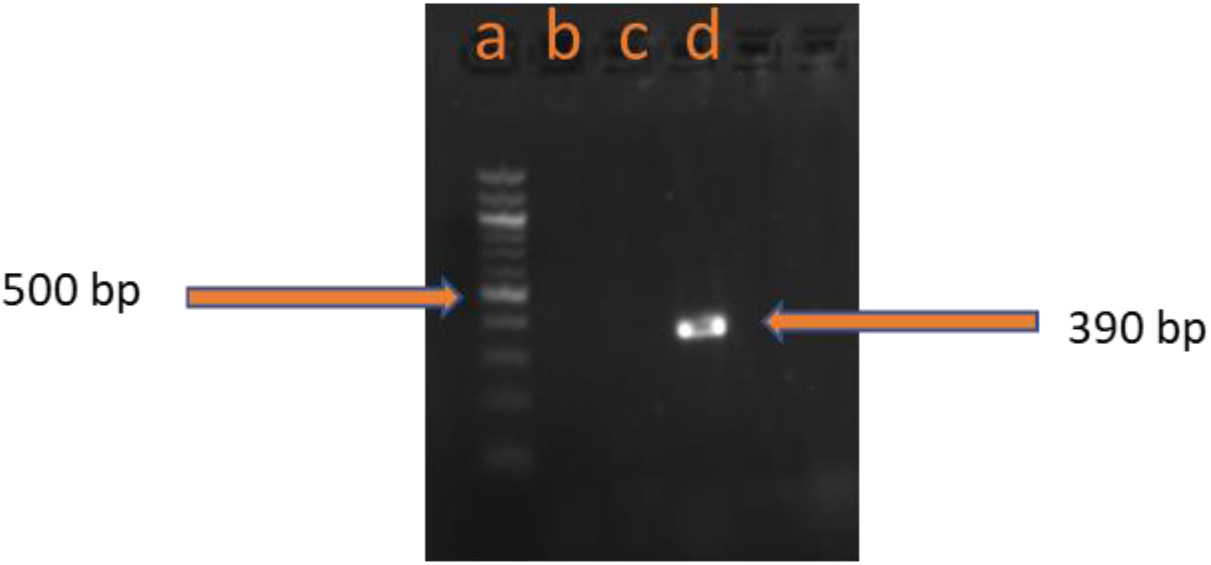
PCR amplification profile of ITS1-5.8S-ITS2 region using ITS1 and ITS4 universal primers, a: 100 bp DNA ladder. d: the amplified fragment for NG06

**Figure 2.**
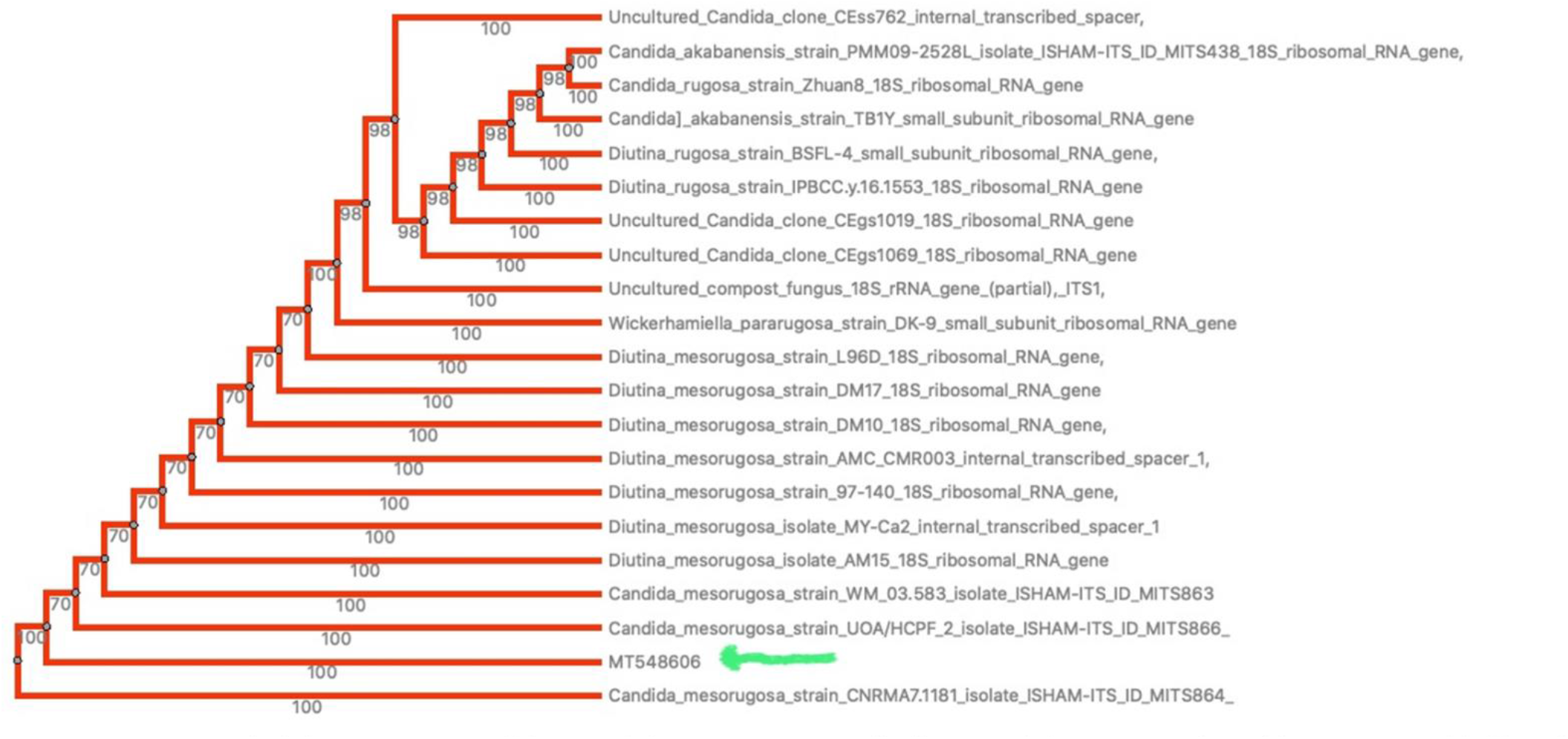
An unrooted phylogenetic tree of the *Candida mesorugosa* and other *Candida* species interred by Maximum-likelihood. The Maximum-likelihood was estimated using Kimura’s two parameter method implemented in Unipro-UGENE version 34.0.

## Discussion

Although *rugosa* complex are relatively common in North India and Latin America, where they account for 18.4% and 1.2% of *Candida* species isolated from patients, respectively, they are rare in clinical samples worldwide (Pfaller et al. 2010; Singh et al. 2011). In this study, a yeast which was germ tube negative in human serum, but morphologically similar to *Candida albicans*, was identified as *Candida mesorugosa* by rDNA targeted sequencing. The *C. mesorugosa* was isolated from a 35-year-old woman who presented with vulvovaginitis after giving birth. The isolate accounts for 2.1% (1/47) of the *Candida* species in our study. This value is above the 0.2% global prevalence of *C. rugosa* complex previously reported (Pfaller et al. 2010). Possibly, *C. rugosa* complex are frequent in clinical yeast isolates in Nigeria but are not detected due to over dependence on phenotypic laboratory methods. A high crude mortality rate (33-83%) is associated with candidemia caused by *C. rugosa* complex (Minces et al. 2009). Moreover, a recent study identified some virulence features, including *C. albicans*-comparable biofilm formation, in *C. mesorugosa* (Montoya et al. 2019). Put together, these call for more surveillance.

## Conclusion

Although the data currently available to us are insufficient to conclude on the causal role of the isolate in the patient’s condition, isolation of a rare yeast known to have poor clinical outcome in severely ill patients calls for attention. This study highlights the need for extensive surveillance of NACS in Nigeria, using very specific methods, such as DNA sequencing.

## Data Availability

All the relevant data are contained in the manuscript

## Acknowledgment

The authors would like to thank the Microbiology Laboratory staff of Ladoke Akintola University of Technology Teaching Hospital, Osogbo for collecting the isolates used in this study.

We would also like to thank Microbiology Laboratory staff of Redeemer’s University, Ede, for providing some technical support for this study.

## Ethical approval

This study was approved by the Research Ethics Committee of Ladoke Akintola University of Technology Teaching Hospital, Osogbo (LTH/EC/2018/04/371).

## Funding

This study was supported by grants from the National Institute of Allergy and Infectious Diseases, NIH-H3Africa (U01HG007480 and U54HG007480 to Dr Christian Happi and a grant from the World Bank (project ACE019 to Redeemer’s University - Dr Christian Happi).

## Author contribution statement

OO and EUD conceived the project. OO, PE, OF and EUD designed the study. OO, JUO and TSO performed the experiments. OO and TA analysed the data. OO prepared the first draft of the manuscript. EUD supervised the study. OF and CH wrote grants applications and secured funding for the study. All authors approved the final version of the manuscript.

## Conflict of interest

The authors declare no conflict of interest

## Notes

### Competing Interest Statement

The authors have declared no competing interest.

### Funding Statement

This study was supported by National Institute of Allergy and Infectious Diseases (NIH-H3Africa U01HG007480 and U54HG007480) and the World Bank (project ACE019).

